# Associations of childhood body composition trajectories and adolescent body dissatisfaction: Longitudinal evidence from the UK Millennium Cohort Study

**DOI:** 10.64898/2025.12.10.25342007

**Authors:** Madelaine Davies Kellock, Francesca Solmi, Anne McMunn, Yvonne Kelly

## Abstract

**Purpose:** Body dissatisfaction is common, particularly among those with higher BMIs. Issues with BMI as an adiposity measure may limit our understanding of body dissatisfaction determinants. We investigate the association of childhood trajectories of BMI, fat mass index (FMI), and fat-free mass index (FFMI) with body dissatisfaction at age 14, and sex differences in these associations.

**Methods:** We used latent class growth analysis to model trajectories of BMI (3-14 years), FMI and FFMI (7-14 years) in Millennium Cohort Study participants, and univariable and multivariable linear regression models to investigate their associations with body dissatisfaction.

**Results:** Among 13,025 participants (49.2% female; 84.4% White), we found three BMI, FMI, and FFMI trajectories (low, moderate, high). Participants with moderate (adjusted mean difference [aMD] 0.32, 95% confidence interval [CI] 0.24, 0.39) and high (aMD 0.71, 95% CI 0.55, 0.86) BMI trajectories and those with moderate (aMD 0.33, 95% CI 0.25, 0.41) and high (aMD 0.73, 95% CI 0.57, 0.89) FMI trajectories had higher body dissatisfaction scores than those with low trajectories. For FFMI trajectories, boys with high (aMD 0.31, 95% CI 0.17, 0.31), but not moderate (aMD 0.07, 95% CI −0.03, 0.17) trajectories had greater body dissatisfaction. In girls, mean differences by FFMI trajectory were consistent with adiposity results.

**Discussion:** Our findings suggest that both boys and girls are susceptible to weight-related body dissatisfaction. Higher FFMI is also associated with body dissatisfaction, but in girls, associations are present at lower levels compared with boys, potentially reflecting higher societal pressures on young girls.

## Introduction

Body dissatisfaction has been rising in young people over the past few decades (1), which is concerning given its association with subsequent mental health problems, including depression and eating disorders (2,3). Through a range of sources from an early age (4,5), young people are exposed to messaging which values some body types and stigmatises others (6). In many Western contexts, thinness in women and lean muscularity in men are seen as socially desirable, while overweight is stigmatised (7). These ideals can become internalised and lead to body dissatisfaction.

Previous cross-sectional research has shown that higher body weight is associated with body dissatisfaction and, consistent with gendered body ideals, there appear to be sex differences in this association (8). In girls, increasing weight is linearly associated with greater body dissatisfaction, while in boys, the association is J-shaped, as both those with low and high weight have higher body dissatisfaction (8). However, prior research has almost exclusively used BMI to indicate weight status as a proxy for adiposity. BMI measures total body weight and cannot distinguish between its component parts, namely fat and fat-free mass. As such, individuals with the same BMI value can have different proportions of fat mass, potentially leading to misclassification of weight status. Some evidence suggests that BMI may be a poor proxy for adiposity, particularly in younger children (9) and in those with non-overweight BMIs, which is the majority of children (10). Fat and fat-free mass may be associated with body dissatisfaction, and their associations with the latter may differ in boys and girls given the social desirability of muscularity in boys.

Few cross-sectional studies have investigated the associations of fat and fat-free mass with body dissatisfaction. These have been predominantly conducted in athletes using body fat percentage to measure adiposity (11). Body fat percentage, however, may be elevated due to high adiposity or low lean mass (12) and so, like BMI, cannot convey the nuanced associations that fat and fat-free mass may have with body dissatisfaction. A small number of cross-sectional studies have utilised fat and fat-free mass measures, with some finding no association with body dissatisfaction for either fat mass or fat-free mass (13,14), and some finding that both higher fat mass (15) and higher fat-free mass are associated with increased body dissatisfaction (15,16). These studies were conducted in athletes and sport science students, who may differ from the general population both in terms of body composition and body dissatisfaction, limiting generalisability. Furthermore, due to their cross-sectional designs, causal inference is limited as individuals experiencing body dissatisfaction may attempt to alter their weight (3).

To our knowledge, only one longitudinal, general population study has investigated the association between fat and fat-free mass and body dissatisfaction (17), using polygenic scores as instrumental variables for body composition, and finding that higher polygenic scores for both fat and fat-free mass are associated with greater body dissatisfaction in adolescence. As this finding contrasts with the theory that higher muscle mass is socially desirable in boys, this association warrants further investigation.

Previous studies have established that there are distinct trajectories of BMI development across childhood (18–20) and have suggested that children with increasing BMI trajectories report higher body dissatisfaction at age 11 (18). There has been little investigation into trajectories of fat and fat-free mass across childhood (21). Using trajectories of body composition, we can disentangle the potentially distinct associations of fat and fat-free mass across childhood and body dissatisfaction in boys and girls, acknowledging the lifecourse nature of weight development.

Therefore, we aimed to investigate whether trajectories of BMI, fat mass index (FMI) and fat-free mass index (FFMI) are differentially associated with body dissatisfaction at age 14, and whether these associations differ by sex. Based on existing knowledge and theory around gendered body ideals, we hypothesised that higher trajectories of adiposity (measured by FMI) across childhood would be associated with greater body dissatisfaction in adolescence, and that the magnitude of association would be greater in girls relative to boys. We also hypothesised that higher trajectories of FFMI, as a proxy for muscularity, would be associated with lower body dissatisfaction in boys only.

## Methods

### Sample

We analysed data from the Millennium Cohort Study, a prospective longitudinal study of children living in the UK who were born between September 2000 and January 2002. The Millennium Cohort Study is a socially and ethnically diverse cohort, with baseline data collection at age 9 months, with subsequent sweeps at ages 3, 5, 7, 11, 14, and 17 years. Ethical review for each sweep was obtained from an NHS Research Ethics Committee, which has been detailed in other reports (22), and written informed consent was obtained from parents and, later, child cohort members themselves. Further details of the cohort, including sampling methods can be found elsewhere (23). We accessed deidentified data on 8^th^ March 2021. For this study, we included participants who had at least one measurement available for each aspect of body composition data up to age 14. In twin pairs, we excluded one twin at random to avoid potential biases arising from shared genetic and environmental factors. As we wanted to compare across multiple measures of body composition, and some were not measured at earlier sweeps, we restricted our analyses to participants with data on all body composition measures (see Exposures).

### Outcome

Body dissatisfaction was measured at age 14 years. In a self-reported questionnaire, participants were asked: “On a scale of 1 to 7, where 1 means completely happy and 7 means not at all happy, how do you feel about the way you look?”. This was recoded to range from 0 to 6, where higher scores indicate greater body dissatisfaction. This item has been widely used as an indicator of body dissatisfaction in prior literature (18,24).

### Exposures

BMI was calculated from measured height and weight at ages 3, 5, 7, 11, and 14 years. Bioelectrical impedance analysis additionally measured body fat percentage at ages 7, 11 and 14 years, from which we calculated FMI and FFMI. Full details of measurement procedures and equations used are provided in the **Supplement** (page 1). Methods used to model trajectories are described in Data analyses.

### Confounders

Potential confounding variables were selected based on existing evidence of their association with weight development and mental health outcomes and were measured when the participants were 9 months of age, unless otherwise specified. These were: child sex at birth and ethnicity, indicators of family socioeconomic position (family income, parent education and occupational class), perinatal factors (child birthweight, maternal pre-pregnancy BMI, breastfeeding status, and maternal use of cigarettes or alcohol during pregnancy), and maternal psychological distress when the child was aged 3 years. We also adjusted for child socioemotional difficulties at age 3, measured by the parent-reported Strengths and Difficulties Questionnaire (SDQ) total score (25). Further detail on confounders can be found in the **Supplement**, page 1).

### Data analyses

All analyses were conducted in Stata 17 (Stata Statistical Software: Release 17. College Station, TX: StataCorp LLC).

#### Trajectory analysis

We modelled trajectories of BMI, FMI and FFMI for children with at least one valid measurement for each of the body composition measures, employing latent class growth analysis (26,27) using maximum likelihood estimation to account for missing data. Starting with a two-class model and increasing the number of classes, we modelled different combinations of polynomial shapes. The optimal polynomial shapes and number of classes was chosen based on goodness of fit (Akaike Information Criterion) and neatness of classification (Entropy Index). These were considered in addition to the ability of each model to identify distinct and meaningful patterns in the data, while maintaining group sample sizes sufficiently large to ensure adequate statistical power for subsequent analyses. After selecting the optimal models, we assigned participants to their most likely trajectory. Full details of latent class trajectory analysis can be found in the **Supplement** (page 3).

#### Descriptive analysis

Among participants with valid exposure data, we described the overlap between trajectories of different body composition measures using proportions. We described characteristics of the overall sample, as well as the distribution of trajectory membership across confounding variables using frequencies and proportions.

#### Missing data

Among participants with complete exposure data, we explored frequencies and proportions of missing data on potential confounders, as well as differences between participants with valid and missing outcome data. Missing data on outcomes and confounders were then imputed using multiple imputation by chained equations, under the assumption that data are missing at random. Fifty datasets were imputed using all analysis variables and additional auxiliary variables to improve precision (see **Supplement**, page 4).

#### Regression analysis

To investigate the association between childhood body composition trajectory and body dissatisfaction at age 14, we used univariable and multivariable linear regression models with separate adjustment for child sex and ethnicity (Model 1), indicators of family socioeconomic position (Model 2), perinatal factors (Model 3), maternal psychological distress (Model 4) and child socioemotional difficulties at age 3 (Model 5). We then ran a fully adjusted model with all confounders in models 1 to 5, before including an exposure*sex interaction to investigate sex differences in the association. If there was evidence of interaction, we reported sex-stratified results.

All regression models were run on a sample with complete exposures with imputed outcomes and confounders. We also included sampling weights to account for the stratified sampling methods.

#### Sensitivity analysis

We ran two sensitivity analyses. First, we repeated unadjusted regression models in the full sample with BMI trajectory data to investigate whether our estimates were impacted by restricting to those with data on both BMI and other body composition measures. Second, we repeated our univariable and multivariable regression analyses in a sample with complete data on exposures, outcomes and confounders.

## Results

### Sample

Of 18,540 participants, 13,025 (70.3%) had valid trajectory group data for all exposures. Approximately half of the participants were male (50.8%), the majority were of White ethnicity (84.4%) and most had a parent educated to degree level or equivalent (42.2%, see **Table 1**).

**Table 1.** Characteristics of sample with valid exposure data and the distribution of trajectory membership among characteristics (N=13025^a^). Proportions of sample with observed data (unweighted)

| Characteristic | N (%) | BMI Trajectory N (%) |  |  | FMI Trajectory N (%) |  |  | FFMI Trajectory N (%) |  |  |
| --- | --- | --- | --- | --- | --- | --- | --- | --- | --- | --- |
|  |  | Low | Moderate | High | Low | Moderate | High | Low | Moderate | High |
| Total (%) |  | 64.3 | 29.0 | 6.7 | 68.6 | 25.2 | 6.2 | 44.9 | 45.9 | 9.2 |
| **Sex at birth** |  |  |  |  |  |  |  |  |  |  |
| Male | 6612 (50.8) | 67.6 | 26.5 | 5.9 | 79.6 | 16.2 | 4.2 | 34.1 | 51.8 | 14.1 |
| Female | 6413 (49.2) | 60.9 | 31.6 | 7.4 | 57.2 | 34.6 | 8.2 | 56.1 | 39.8 | 4.1 |
| **Ethnicity** |  |  |  |  |  |  |  |  |  |  |
| White | 10968 (84.4) | 64.6 | 29.2 | 6.2 | 69.8 | 24.6 | 5.6 | 43.4 | 47.2 | 9.4 |
| Mixed | 346 (2.7) | 65.6 | 25.4 | 9.0 | 67.3 | 23.7 | 9.0 | 46.0 | 44.2 | 9.8 |
| Indian | 312 (2.4) | 68.6 | 26.0 | 5.4 | 64.1 | 27.9 | 8.0 | 62.8 | 34.6 | 2.6 |
| Pakistani or Bangladeshi | 828 (6.4) | 62.2 | 27.9 | 9.9 | 59.5 | 30.6 | 9.9 | 58.3 | 35.0 | 6.7 |
| Black or Black British | 383 (2.9) | 52.0 | 36.0 | 12.0 | 57.7 | 30.6 | 11.7 | 37.9 | 47.2 | 14.9 |
| Other, including Chinese and Arab | 160 (1.2) | 70.0 | 23.1 | 6.9 | 67.5 | 26.9 | 5.6 | 57.5 | 36.9 | 5.6 |
| **Family income at 9 months** |  |  |  |  |  |  |  |  |  |  |
| Most disadvantaged | 2804 (21.6) | 60.4 | 30.3 | 9.3 | 63.8 | 27.1 | 9.1 | 43.9 | 45.5 | 10.6 |
| Second | 2786 (21.4) | 62.4 | 30.0 | 7.6 | 65.5 | 27.4 | 7.1 | 44.5 | 45.2 | 10.3 |
| Third | 2526 (19.4) | 62.4 | 31.1 | 6.5 | 67.3 | 26.5 | 6.2 | 43.0 | 47.5 | 9.5 |
| Fourth | 2521 (19.4) | 64.7 | 29.7 | 5.6 | 70.0 | 25.3 | 4.7 | 43.6 | 47.7 | 8.7 |
| Least disadvantaged | 2355 (18.1) | 72.8 | 23.5 | 3.7 | 77.5 | 19.2 | 3.3 | 49.9 | 43.6 | 6.5 |
| **Highest parental education level at 9 months** |  |  |  |  |  |  |  |  |  |  |
| None or overseas | 1403 (10.8) | 60.1 | 29.9 | 10.0 | 62.3 | 28.2 | 9.5 | 47.2 | 41.8 | 11.0 |
| NVQ Level 1 | 775 (5.9) | 62.6 | 29.9 | 7.5 | 67.6 | 25.7 | 6.7 | 46.2 | 43.6 | 10.2 |
| NVQ Level 2 | 3199 (24.6) | 60.6 | 31.4 | 8.0 | 64.5 | 27.7 | 7.8 | 42.0 | 47.8 | 10.2 |
| NVQ Level 3 | 2151 (16.5) | 63.6 | 29.2 | 7.2 | 68.3 | 25.5 | 6.2 | 44.2 | 45.3 | 10.5 |
| NVQ Level 4 | 4593 (35.3) | 67.2 | 27.9 | 4.9 | 71.8 | 23.5 | 4.7 | 45.2 | 47.2 | 7.6 |
| NVQ Level 5 | 896 (6.9) | 72.5 | 23.9 | 3.6 | 77.5 | 19.7 | 2.8 | 50.9 | 42.2 | 6.9 |
| **Highest parental occupational class at 9 months** |  |  |  |  |  |  |  |  |  |  |
| Not in work | 2924 (22.5) | 60.5 | 30.6 | 8.9 | 64.2 | 26.9 | 8.9 | 44.0 | 45.0 | 10.9 |
| Routine or semi-routine | 1900 (14.6) | 60.7 | 30.5 | 8.8 | 63.6 | 28.3 | 8.1 | 42.8 | 46.6 | 10.6 |
| Lower supervisory or technical | 1043 (8.0) | 59.2 | 33.2 | 7.6 | 64.6 | 27.9 | 7.5 | 41.6 | 48.4 | 10.0 |
| Small employers or self-employed | 863 (6.7) | 63.6 | 29.8 | 6.6 | 67.3 | 26.4 | 6.3 | 44.8 | 45.4 | 9.7 |
| Intermediate occupations | 1199 (9.2) | 66.3 | 27.5 | 6.2 | 70.1 | 24.8 | 5.2 | 45.8 | 45.9 | 8.3 |
| Managerial or professional | 5050 (38.9) | 68.5 | 27.1 | 4.4 | 73.6 | 22.6 | 3.8 | 46.8 | 45.6 | 7.6 |
| **Birthweight** |  |  |  |  |  |  |  |  |  |  |
|  |  | Low | Moderate | High | Low | Moderate | High | Low | Moderate | High |
| Low birthweight ($\leq 2.5$kg) | 890 (6.9) | 68.7 | 24.9 | 6.4 | 67.9 | 24.8 | 7.3 | 55.8 | 36.5 | 7.6 |
| Birthweight $> 2.5$kg | 12106 (93.1) | 64.0 | 29.3 | 6.7 | 68.6 | 25.3 | 6.1 | 44.1 | 46.6 | 9.3 |
| **Maternal pre-pregnancy BMI** |  |  |  |  |  |  |  |  |  |  |
| BMI $< 25$ | 8416 (70.3) | 71.4 | 24.6 | 4.0 | 74.8 | 21.5 | 3.7 | 50.4 | 43.2 | 6.4 |
| BMI 25 – 29.9 | 2465 (20.6) | 53.6 | 36.7 | 9.7 | 60.0 | 31.2 | 8.8 | 35.6 | 51.6 | 12.8 |
| BMI $\geq 30$ | 1083 (9.1) | 38.2 | 43.1 | 18.7 | 45.9 | 37.2 | 16.9 | 24.3 | 53.6 | 22.1 |
| **Breastfed** |  |  |  |  |  |  |  |  |  |  |
| No | 3870 (29.8) | 60.8 | 31.0 | 8.2 | 64.7 | 27.4 | 7.9 | 42.5 | 47.2 | 10.3 |
| Yes | 9135 (70.2) | 65.8 | 28.2 | 6.0 | 70.2 | 24.3 | 5.5 | 46.0 | 45.3 | 8.7 |
| **Cigarette use in pregnancy** |  |  |  |  |  |  |  |  |  |  |
| None | 9455 (72.6) | 65.8 | 28.3 | 5.9 | 69.4 | 24.9 | 5.7 | 46.5 | 45.2 | 8.4 |
| Any | 3564 (27.4) | 60.5 | 31.0 | 8.5 | 63.8 | 27.2 | 9.0 | 36.4 | 49.8 | 13.8 |
| **Alcohol use in pregnancy** |  |  |  |  |  |  |  |  |  |  |
| None | 9020 (69.4) | 63.0 | 29.7 | 7.3 | 67.0 | 26.0 | 7.0 | 45.1 | 45.3 | 9.6 |
| Any | 3982 (30.6) | 67.4 | 27.3 | 5.3 | 72.1 | 23.5 | 4.3 | 44.6 | 47.1 | 8.3 |
| **Maternal psychological distress at 3 years** |  |  |  |  |  |  |  |  |  |  |
| Low ($< 5$) | 8591 (75.0) | 67.0 | 27.1 | 5.9 | 69.6 | 24.8 | 5.6 | 44.3 | 46.8 | 8.9 |
| Moderate (5-11) | 2472 (21.6) | 65.2 | 27.2 | 7.6 | 66.5 | 26.3 | 7.2 | 42.4 | 46.8 | 10.8 |
| Severe ($\geq 12$) | 388 (3.4) | 65.5 | 26.0 | 8.6 | 65.7 | 26.0 | 8.3 | 43.0 | 47.4 | 9.5 |
| **Child total SDQ score at 3 years** |  |  |  |  |  |  |  |  |  |  |
| Close to average ($\leq 13$) | 9591 (79.0) | 64.8 | 29.1 | 6.1 | 69.2 | 25.0 | 5.8 | 45.3 | 46.0 | 8.7 |
| Slightly raised (14-16) | 1276 (10.5) | 61.4 | 30.5 | 8.1 | 67.0 | 26.3 | 6.7 | 40.8 | 48.4 | 10.7 |
| High (17-19) | 678 (5.6) | 63.1 | 28.9 | 8.0 | 68.6 | 23.7 | 7.7 | 42.6 | 45.1 | 12.3 |
| Very high ($\geq 20$) | 593 (4.9) | 61.9 | 28.7 | 9.4 | 67.6 | 24.0 | 8.4 | 44.2 | 44.3 | 11.5 |

a Ns may not total 13025 due to missing data; see Supplementary Information for more on missing data.

### Body composition trajectories

For each measure of body composition, the three-trajectory model, including a low, a moderate, and a high trajectory group (**Fig 1**) was considered optimal. Full results of trajectory modelling can be found in the **Supplement** (page 5). The majority of participants were assigned to a low trajectory of BMI (64.3%) and FMI (68.6%), with 29.0% (BMI) and 25.2% (FMI) of participants assigned to a moderate trajectory and 6.7% (BMI) and 6.2% (FMI) to a high trajectory (**Table 1**). Roughly equal proportions of participants had low and moderate trajectories of FFMI (44.9% and 45.9%, respectively), while 9.2% had high trajectories. There was large overlap between BMI and adiposity trajectory membership, but less so for FFMI (**Table 2**).

**Fig 1.**
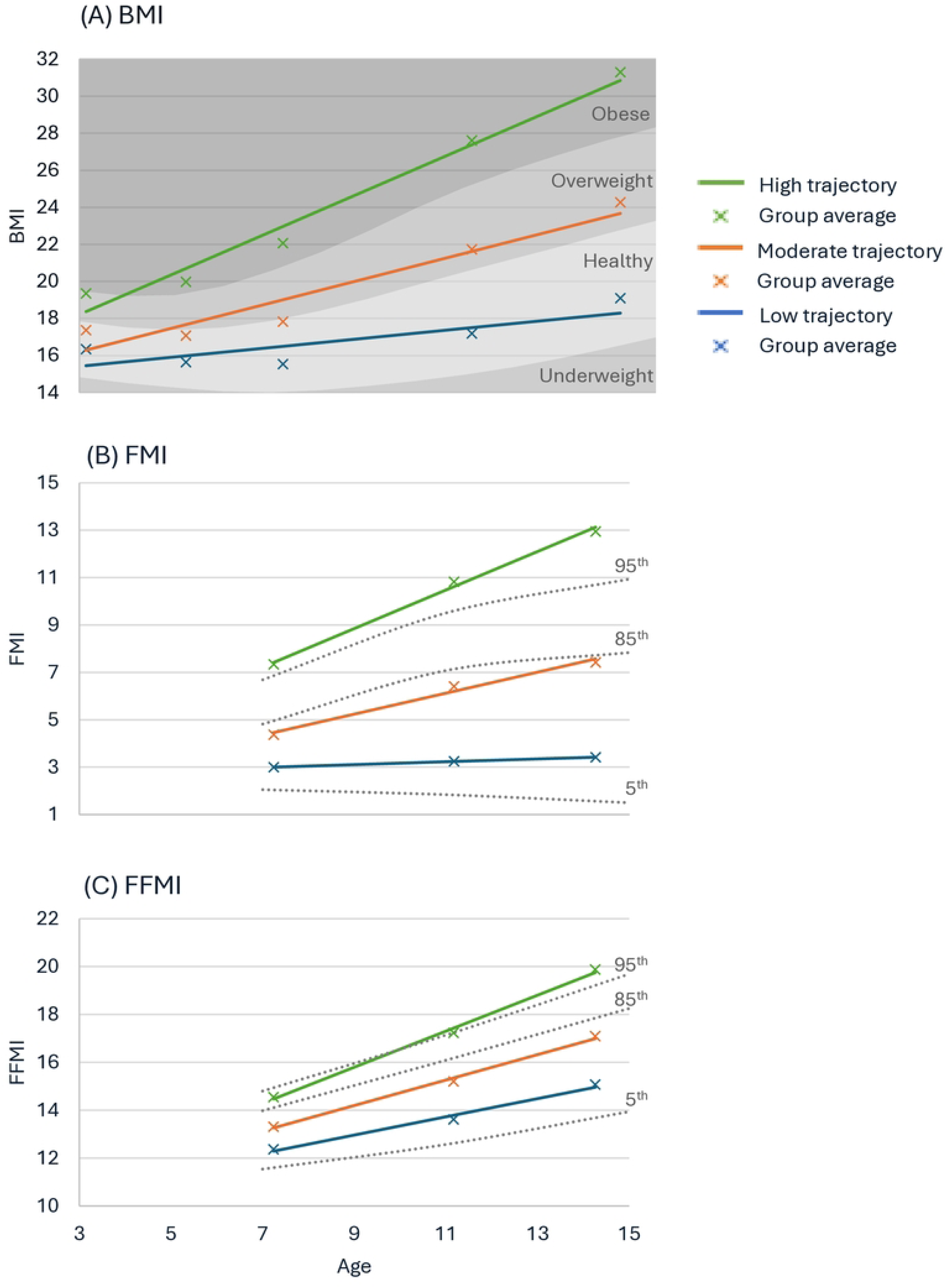
Body composition trajectories for (A) BMI, (B) FMI, (C) FFMI. Crosses show average data points for each group with lines representing the estimated trajectories. For BMI, shaded areas show the International Obesity Task Force thresholds for categorising child BMI. For FMI and FFMI dashed lines reflect centiles within the MCS sample.

**Table 2.**
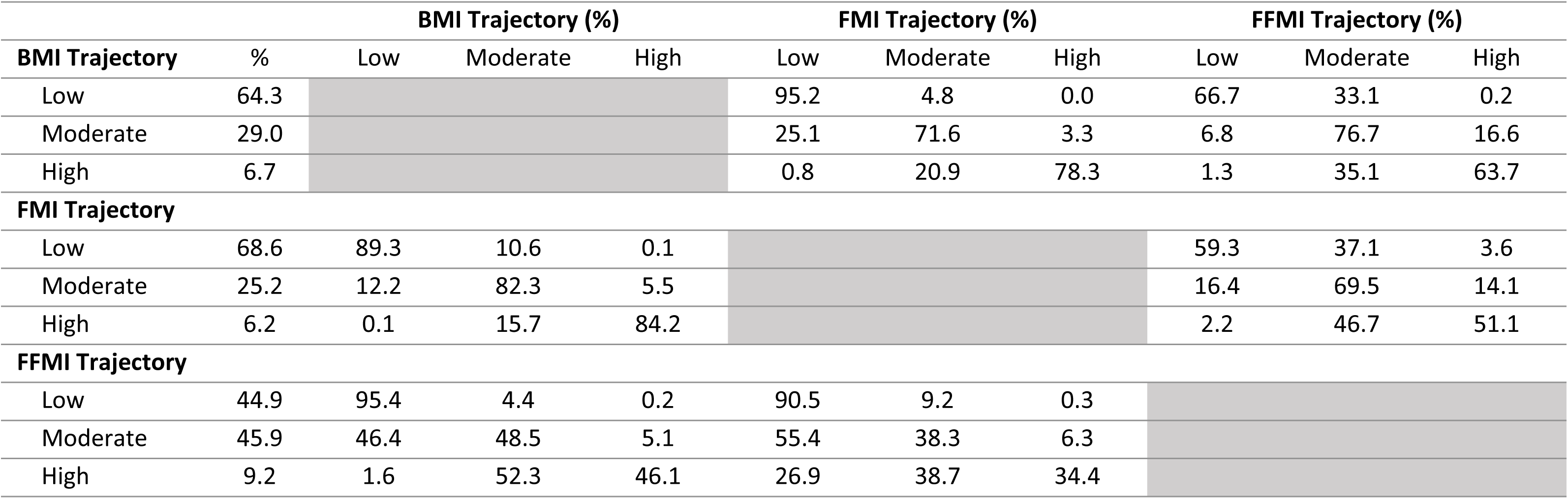
Overlap between body composition trajectory membership.

A greater proportion of girls, participants of mixed, Pakistani, Bangladeshi, or Black ethnicity, and from more socioeconomically disadvantaged backgrounds had higher trajectories of BMI and FMI across childhood (**Table 1**). Higher maternal pre-pregnancy BMI, psychological distress, and participant SDQ score at age 3 were all associated with higher participant BMI and FMI trajectories. In contrast to these exposures, a greater proportion of boys and participants from more socioeconomically advantaged backgrounds had higher trajectories of FFMI (**Table 1**).

### Missing data

Among those with valid exposure data, 5,092 were missing data on outcomes or confounders. A greater proportion of those with missing data were male, from ethnic minority backgrounds, more socioeconomically disadvantaged, and had greater socioemotional difficulties at age 3 as well as mothers with higher levels of psychological distress (see **Supplement**, page 12).

### Association between childhood body composition trajectories and body satisfaction at age 14

In unadjusted analyses (**Table 3**), there was evidence that children with moderate (mean difference [MD] 0.41, 95% Confidence Intervals [CI] 0.34, 0.49) and high (MD 0.87, 95% CI 0.71, 1.02) trajectories of BMI had higher body dissatisfaction scores in adolescence, compared to those with low trajectories. Patterns of association were similar for FMI, though mean differences were larger for both moderate (MD 0.59, 95% CI 0.51, 0.67) and high (MD 1.02, 95% CI 0.85, 1.18) trajectories. In fully adjusted models, there remained strong evidence of associations for both BMI (moderate: adjusted MD 0.32, 95% CI 0.24, 0.39; high: aMD 0.71, 95% CI 0.55, 0.86) and FMI (moderate: aMD 0.33, 95% CI 0.25, 0.41; high: aMD 0.73, 95% CI 0.57, 0.89), and mean differences were consistent across the two measures. There was no evidence of sex differences in the association of BMI (interaction test p value for moderate trajectory=0.216; high p=0.140) or FMI (moderate p=0.968; high p=0.207) and body dissatisfaction.

**Table 3.** Results of univariable and multivariable linear regression analysis of the association between body composition trajectory and body dissatisfaction at age 14 in participants with valid exposure data and imputed outcomes and confounders (sampling weights applied).

| Model | *Mean difference in body dissatisfaction at age 14 years (95% confidence interval)* |  |  |  |  |  |  |  |  |
| --- | --- | --- | --- | --- | --- | --- | --- | --- | --- |
|  | BMI Trajectory |  |  | FMI Trajectory |  |  | FFMI Trajectory |  |  |
|  | Low | Moderate | High | Low | Moderate | High | Low | Moderate | High |
| Model 0<br>(unadjusted) | ref | 0.41<br>(0.34, 0.49) | 0.87<br>(0.71, 1.02) | ref | 0.59<br>(0.51, 0.67) | 1.02<br>(0.85, 1.18) | ref | 0.07<br>(-0.00, 0.15) | 0.17<br>(0.03, 0.30) |
| Model 1<br>(0 + child characteristics<sup>a</sup>) | ref | 0.36<br>(0.28, 0.43) | 0.80<br>(0.66, 0.96) | ref | 0.38<br>(0.30, 0.46) | 0.83<br>(0.68, 1.00) | ref | 0.27<br>(0.19, 0.34) | 0.59<br>(0.45, 0.72) |
| Model 2<br>(0 + family SEP<sup>b</sup>) | ref | 0.40<br>(0.33, 0.48) | 0.84<br>(0.68, 1.00) | ref | 0.58<br>(0.50, 0.66) | 0.99<br>(0.83, 1.15) | ref | 0.06<br>(- 0.01, 0.14) | 0.14<br>(0.01, 0.27) |
| Model 3<br>(0 + perinatal factors<sup>c</sup>) | ref | 0.40<br>(0.31, 0.47) | 0.80<br>(0.64, 0.96) | ref | 0.56<br>(0.48, 0.65) | 0.96<br>(0.80, 1.12) | ref | 0.04<br>(-0.04, 0.11) | 0.07<br>(-0.07, 0.20) |
| Model 4<br>(0 + maternal psychological distress<sup>d</sup>) | ref | 0.41<br>(0.34, 0.49) | 0.85<br>(0.69, 1.01) | ref | 0.58<br>(0.51, 0.66) | 1.00<br>(0.84, 1.17) | ref | 0.07<br>(-0.00, 0.15) | 0.16<br>(0.03, 0.29) |
| Model 5<br>(0 + child socioemotional difficulties<sup>e</sup>) | ref | 0.41<br>(0.34, 0.49) | 0.86<br>(0.70, 1.01) | ref | 0.59<br>(0.51, 0.67) | 1.01<br>(0.85, 1.17) | ref | 0.07<br>(-0.01, 0.14) | 0.15<br>(0.02, 0.28) |
| Model 6<br>(0 + all confounders in Models 1-5) | ref | 0.32<br>(0.24, 0.39) | 0.71<br>(0.55, 0.86) | ref | 0.33<br>(0.25, 0.41) | 0.73<br>(0.57, 0.89) | ref | 0.23<br>(0.15, 0.30) | 0.48<br>(0.34, 0.62) |
| Exposure × sex interaction<br>p value | ref | 0.216 | 0.140 | ref | 0.968 | 0.207 | ref | <0.001 | 0.003 |
| Mean difference in boys<br>(N=6612) | ref | - | - | ref | - | - | ref | 0.07<br>(-0.03, 0.17) | 0.31<br>(0.17, 0.46) |
| Mean difference in girls<br>(N=6413) | ref | - | - | ref | - | - | ref | 0.36<br>(0.26, 0.47) | 0.79<br>(0.47, 1.10) |

<sup>a</sup> Child sex at birth, ethnicity.
<sup>b</sup> Family income quintile, parental educational qualification level, parental occupational class.
<sup>c</sup> Child birthweight, maternal pre-pregnancy BMI, breastfeeding, cigarette and alcohol use in pregnancy.
<sup>d</sup> Maternal psychological distress score at 3 years old.
<sup>e</sup> Child total SDQ score at 3 years old.

In contrast, for FFMI trajectories, there was evidence in unadjusted models of smaller mean differences in body dissatisfaction among those with moderate (MD 0.07, 95% CI −0.00, 0.15) and high FFMI (MD 0.17, 95% CI 0.03, 0.30) compared to the low trajectory group (**Table 3**). When adjusting for child sex and ethnicity, the magnitude of these mean differences increased (moderate: aMD 0.27, 95% CI 0.19, 0.34; high: aMD 0.59, 95% CI 0.45, 0.72), such that these were more consistent with associations seen for BMI and FMI trajectories. In the fully adjusted model, there was strong evidence that participants with moderate (aMD 0.23, 95% CI 0.15, 0.30) and high (aMD 0.48, 95% CI 0.34, 0.62) FFMI trajectories had higher body dissatisfaction than those with low FFMI trajectories, but mean differences were smaller than those seen for adiposity measures. There was strong evidence that these associations differed by sex (interaction test p value for moderate trajectory <0.001; high p=0.003).

In boys, only those with high FFMI trajectories had greater body dissatisfaction (moderate: aMD 0.07, 95% CI −0.03, 0.17; high: aMD 0.31, 95% CI 0.17, 0.46). In girls, the association was consistent with those seen for BMI and FMI (moderate FFMI: aMD 0.36, 95% CI 0.36, 0.47; high: aMD 0.79, 95% CI 0.47, 1.10).

### Sensitivity analyses

When we repeated regression models in a sample of participants with valid BMI trajectory data (N=14,765), results were consistent with those of the main analysis. Similarly, when we repeated our analyses in a sample with complete data on exposures, outcomes and confounders, we found that associations and results of interaction tests were similar to those in the main analysis. See **Supplement** (page 16) for full results.

## Discussion

In this study we found distinct trajectories of BMI, FMI and FFMI. Our results suggest that higher childhood trajectories of adiposity, measured by BMI or FMI, were associated with high levels of body dissatisfaction in adolescence. There was no evidence that this association differed by sex, which was in contrast to our initial hypothesis. However, we did find evidence that the association between FFMI trajectory and body dissatisfaction differed by sex. We had hypothesised that a negative association would be observed in boys due to the social desirability of muscularity; instead, we found that boys with high FFMI trajectories had greater body dissatisfaction than those with low FFMI, but mean differences were smaller than adiposity estimates. In girls, the association between FFMI and body dissatisfaction was consistent with adiposity measures.

### Limitations

Body composition data was measured using bioelectrical impedance analysis, rather than the gold standard dual x-ray absorptiometry scans. Studies have previously shown concordance between the two methods (28,29). However, this limited our investigation of distinct components of body composition to fat and fat-free mass. We were unable to further break down these components, for instance into lean mass, which could be measured in dual x-ray absorptiometry scans. In this study, fat-free mass is used as a proxy for lean mass, but factors other than lean mass may contribute to raised fat-free mass, such as elevated bone mass (30).

We created indices of body composition measures by dividing each indicator by height squared, as is standard practice; but recent evidence suggests that this may not sufficiently account for the correlation between weight and height, particularly when using fat and fat-free mass (31). Therefore, there may be residual confounding by height, and this could differentially impact girls and boys as taller stature is more socially desirable in men. Further, although we adjusted for a wide range of child and family characteristics, there may be residual confounding, for instance by unmeasured factors within the family environment such as parental eating disorders and attitudes to weight.

### Interpretation of findings

Based on theory of the gendered nature of sociocultural body ideals, we had expected to find that higher adiposity in girls and lower muscularity in boys are associated with greater body dissatisfaction. Instead, we found that high muscularity, as measured by FFMI, was associated with greater body dissatisfaction in both sexes and that the magnitude of this association was greater in girls than in boys. It is possible that individuals with higher FFMI trajectories could be those highly engaged across childhood in sports where participants have a higher prevalence of eating disorders and body image concerns, such as those where leanness is considered a competitive advantage, e.g., gymnastics and swimming (32). Alternatively, previous research has shown that with increases in adiposity, there occur adaptive increases in lean mass (33), and therefore the high FFMI trajectory group could include both individuals with lean muscular body types as well as those with high adiposity levels or elevated bone mass. This may explain why boys with high FFMI trajectories have higher dissatisfaction, in contrast with our initial hypothesis. Nevertheless, the positive association we observed between FFMI and body dissatisfaction in both sexes is consistent with a previous general population study in adolescents born ten years earlier (17). Furthermore, we observed associations with body dissatisfaction in both moderate and high FFMI levels in girls, but only for high FFMI in boys. This suggests that body dissatisfaction may occur at smaller body sizes in girls compared to boys, potentially reflecting higher societal pressures on girls.

We did not find evidence that the association between adiposity trajectory and body dissatisfaction differed by sex. This is somewhat consistent with previous cross-sectional studies using BMI, which have shown that sex differences are present at lower BMI values, but both girls and boys with higher BMIs are more dissatisfied (8). Young people are exposed to weight stigma from many sources, including in interpersonal relationships and via various forms of media (6,34). Additionally, public health strategies aimed at reducing childhood overweight often target primary school-aged children. The statutory primary school curriculum in England includes learning about the risks of obesity and an understanding of calories and other nutritional content (35). The National Child Measurement Programme has also seen the introduction of children being weighed in school in the first and final years of primary school (36), and a report from a recent Select Committee inquiry into body image called for annual health assessments in schools including weight measurements (37), implying that by identifying and reducing overweight, body image concerns may be prevented. It is possible that these strategies’ focus on weight could contribute to increased stigma and lead to feelings of guilt and shame in children with higher weights.

## Conclusion

Body dissatisfaction is often considered to only affect women and girls, yet it has been rising in adolescent boys (1,38). Our findings suggest that both girls and boys with higher childhood adiposity are at risk of body dissatisfaction in adolescence. This is concerning as body dissatisfaction is a risk factor for mental health problems (2,3) and is likely to contribute to increased weight gain over time (39). These findings invite increased awareness of body dissatisfaction in boys and action to reduce young people’s exposure to weight-stigmatised messaging.

## Data Availability

Millennium Cohort Study data are publicly available free of charge through the UK Data Service. Further information about the data can be found on the Centre for Longitudinal Studies website: https://cls.ucl.ac.uk/cls-studies/millennium-cohort-study/

## Acknowledgements

We are grateful to the Centre for Longitudinal Studies (CLS) for use of these data, and to the UK Data Service for making these freely available. The Economic and Social Research Council (ESRC) funds the CLS Resource Centre [ES/W013142/1] which provides core support for the CLS cohort studies. While the CLS Resource Centre makes these data available, CLS does not bear any responsibility for the analysis or interpretation of these data by researchers. The CLS cohorts are only possible due to the commitment and enthusiasm of their participants; their time and contributions are gratefully acknowledged.

## Supporting Information

S1 Appendix.

## References

1. The Children’s Society. The Good Childhood Report 2023 [Internet]. The Children’s Society; 2023. Available from: https://www.childrenssociety.org.uk/sites/default/files/2023-09/The%20Good%20Childhood%20Report%202023.pdf

2. Bornioli A, Lewis-Smith H, Slater A, Bray I. Body dissatisfaction predicts the onset of depression among adolescent females and males: a prospective study. J Epidemiol Community Health. 2021 Apr 1;75(4):343–8.

3. Sharpe H, Griffiths S, Choo TH, Eisenberg ME, Mitchison D, Wall M, et al. The relative importance of dissatisfaction, overvaluation and preoccupation with weight and shape for predicting onset of disordered eating behaviors and depressive symptoms over 15 years. Int J Eat Disord. 2018;51(10):1168–75.

4. Harrison S, Rowlinson M, Hill AJ. “No fat friend of mine”: Young children’s responses to overweight and disability. Body Image. 2016 Sep 1;18:65–73.

5. Shroff H, Thompson JK. The tripartite influence model of body image and eating disturbance: A replication with adolescent girls. Body Image. 2006 Mar;3(1):17–23.

6. Brown A, Flint SW, Batterham RL. Pervasiveness, impact and implications of weight stigma. eClinicalMedicine [Internet]. 2022 May 1 [cited 2023 Apr 17];47. Available from: https://www.thelancet.com/journals/eclinm/article/PIIS2589-5370(22)00138-9/fulltext

7. Smolak L, Murnen SK. Drive for leanness: Assessment and relationship to gender, gender role and objectification. Body Image. 2008 Sep 1;5(3):251–60.

8. Austin SB, Haines J, Veugelers PJ. Body satisfaction and body weight: gender differences and sociodemographic determinants. BMC Public Health. 2009 Aug 27;9:313.

9. Vanderwall C, Randall Clark R, Eickhoff J, Carrel AL. BMI is a poor predictor of adiposity in young overweight and obese children. BMC Pediatr. 2017 Jun 2;17:135.

10. Freedman DS, Wang J, Maynard LM, Thornton JC, Mei Z, Pierson RN, et al. Relation of BMI to fat and fat-free mass among children and adolescents. Int J Obes 2005. 2005 Jan;29(1):1–8.

11. Webb MD, Melough MM, Earthman CP, Katz SE, Pacanowski CR. Associations between anthropometry, body composition, and body image in athletes: a systematic review. Front Psychol [Internet]. 2024 May 13 [cited 2025 Mar 6];15. Available from: https://www.frontiersin.org/journals/psychology/articles/10.3389/fpsyg.2024.1372331/full

12. Wells JCK. Toward body composition reference data for infants, children, and adolescents. Adv Nutr Bethesda Md. 2014 May;5(3):320S–9S.

13. Kagawa M, Iwamoto S, Ishikawa-Takata K, Ota M. Physical Characteristics and Body Image of Japanese Female University Long-Distance Runners. Appl Sci. 2023 Jan;13(11):6442.

14. Remmel L, Jürimäe J, Tamm AL, Purge P, Tillmann V. The Associations of Body Image Perception with Serum Resistin Levels in Highly Trained Adolescent Estonian Rhythmic Gymnasts. Nutrients. 2021 Sep;13(9):3147.

15. Gualdi-Russo E, Rinaldo N, Masotti S, Bramanti B, Zaccagni L. Sex Differences in Body Image Perception and Ideals: Analysis of Possible Determinants. Int J Environ Res Public Health. 2022 Jan;19(5):2745.

16. Hernández-Martínez A, González-Martí I, Jordán ORC. Detection of Muscle Dysmorphia symptoms in male weightlifters. An Psicol Ann Psychol. 2017;33(1):204–10.

17. Abdulkadir M, Hubel C, Herle M, Loos RJ, Breen G, Bulik CM, et al. Eating disorder symptoms and their associations with anthropometric and psychiatric polygenic scores. Eur Eat Disord Rev J Eat Disord Assoc. 2022;30(3):221–36.

18. Kelly Y, Patalay P, Montgomery S, Sacker A. BMI Development and Early Adolescent Psychosocial Well-Being: UK Millennium Cohort Study. Pediatrics. 2016 Dec;138(6):e20160967.

19. Pryor LE, Tremblay RE, Boivin M, Touchette E, Dubois L, Genolini C, et al. Developmental trajectories of body mass index in early childhood and their risk factors: an 8-year longitudinal study. Arch Pediatr Adolesc Med. 2011 Oct;165(10):906–12.

20. Garden FL, Marks GB, Simpson JM, Webb KL. Body Mass Index (BMI) Trajectories from Birth to 11.5 Years: Relation to Early Life Food Intake. Nutrients. 2012 Oct 9;4(10):1382–98.

21. Lovinsky-Desir S, Lussier SJ, Calatroni A, Gergen PJ, Rivera-Spoljaric K, Bacharier LB, et al. Trajectories of adiposity indicators and association with asthma and lung function in urban minority children. J Allergy Clin Immunol. 2021 Nov 1;148(5):1219–1226.e7.

22. Shepherd P, Gilbert E. Millennium Cohort Study: Ethical Review and Consent [Internet]. Centre for Longitudinal Studies; 2019. Available from: https://cls.ucl.ac.uk/wp-content/uploads/2017/07/MCS-Ethical-Approval-and-Consent-2019.pdf?_gl=1*1×3xykn*_up*MQ..*_ga*MTcwODAxNzg5MS4xNzYzNDY3Njg4*_ga_EYRQV4V0KV*czE3NjM0Njc2ODckbzEkZzAkdDE3NjM0Njc2ODckajYwJGwwJGgw

23. Connelly, Roxanne, Platt, Lucinda. Cohort Profile: UK Millennium Cohort Study (MCS). Int J Epidemiol. 2014;43(6):1719–25.

24. Blundell E, Stavola BLD, Kellock MD, Kelly Y, Lewis G, McMunn A, et al. Longitudinal pathways between childhood BMI, body dissatisfaction, and adolescent depression: an observational study using the UK Millenium Cohort Study. Lancet Psychiatry. 2024 Jan 1;11(1):47–55.

25. Goodman R. Psychometric properties of the strengths and difficulties questionnaire. J Am Acad Child Adolesc Psychiatry. 2001 Nov;40(11):1337–45.

26. Jones BL, Nagin DS. A Note on a Stata Plugin for Estimating Group-based Trajectory Models. Sociol Methods Res. 2013 Nov;42(4):608–13.

27. Jung T, Wickrama K a. S. An Introduction to Latent Class Growth Analysis and Growth Mixture Modeling. Soc Personal Psychol Compass. 2008;2(1):302–17.

28. Meredith-Jones KA, Williams SM, Taylor RW. Bioelectrical impedance as a measure of change in body composition in young children. Pediatr Obes. 2015;10(4):252–9.

29. Barreira TV, Staiano AE, Katzmarzyk PT. Validity assessment of a portable bioimpedance scale to estimate body fat percentage in White and African–American children and adolescents. Pediatr Obes. 2013;8(2):e29–32.

30. Howe LD, Lawlor DA, Propper C. Trajectories of socioeconomic inequalities in health, behaviours and academic achievement across childhood and adolescence. J Epidemiol Community Health. 2013 Apr 1;67(4):358–64.

31. Bridger Staatz C, Kelly Y, Lacey RE, Hardy R. Area-level and family-level socioeconomic position and body composition trajectories: longitudinal analysis of the UK Millennium Cohort Study. Lancet Public Health. 2021;6(8):e598–607.

32. Mancine RP, Gusfa DW, Moshrefi A, Kennedy SF. Prevalence of disordered eating in athletes categorized by emphasis on leanness and activity type – a systematic review. J Eat Disord. 2020 Sep 29;8(1):47.

33. Cooper R, Hardy R, Bann D, Aihie Sayer A, Ward KA, Adams JE, et al. Body Mass Index From Age 15 Years Onwards and Muscle Mass, Strength, and Quality in Early Old Age: Findings From the MRC National Survey of Health and Development. J Gerontol A Biol Sci Med Sci. 2014 Oct;69(10):1253–9.

34. Eisenberg ME, Carlson-McGuire A, Gollust SE, Neumark-Sztainer D. A Content Analysis of Weight Stigmatization in Popular Television Programming for Adolescents. Int J Eat Disord. 2015 Sep;48(6):759–66.

35. Department for Education. Statutory guidance: Physical health and mental wellbeing (Primary and secondary) [Internet]. Department for Education; 2021. Available from: https://www.gov.uk/government/publications/relationships-education-relationships-and-sex-education-rse-and-health-education/physical-health-and-mental-wellbeing-primary-and-secondary

36. Department of Health & Social Care. Childhood obesity: a plan for action, Chapter 2 [Internet]. Department of Health and Social Care; 2018. Available from: https://assets.publishing.service.gov.uk/media/5b30a40de5274a55c78cef32/childhood-obesity-a-plan-for-action-chapter-2.pdf

37. Health and Social Care Committee. The impact of body image on mental and physical health: Second Report of Session 2022–23 [Internet]. Health and Social Care Committee; 2022. Available from: https://publications.parliament.uk/pa/cm5803/cmselect/cmhealth/114/report.html

38. Solmi F, Sharpe PhD H, Gage SH, Maddock J, Lewis G, Patalay P. Changes in the Prevalence and Correlates of Weight-Control Behaviors and Weight Perception in Adolescents in the UK, 1986-2015. JAMA Pediatr. 2021 Mar 1;175(3):267–75.

39. Yoon C, Mason SM, Hooper L, Eisenberg ME, Neumark-Sztainer D. Disordered Eating Behaviors and 15-year Trajectories in Body Mass Index: Findings From Project Eating and Activity in Teens and Young Adults (EAT). J Adolesc Health Off Publ Soc Adolesc Med. 2020 Feb;66(2):181–8.

